# Plasmalogens and their Associations with Brain Function and Structure in Older Community Dwelling African Americans

**DOI:** 10.64898/2026.03.26.26349429

**Authors:** AA Weaver, RC Shah, L Du, V Senanayake, DB Goodenowe, LL Barnes

## Abstract

**BACKGROUND:** Recent studies consisting primarily of white participants have found lowered plasmalogen levels to be associated with lower cognitive function. We explore the association of blood plasmalogen levels with global cognition and brain imaging metrics in older African Americans.

**METHODS:** Included in these cross-sectional analyses were participants in the Minority Aging Research Study (MARS) and the Rush Clinical Core without dementia, available serum lipid levels, and a concurrent cognitive function assessment. A plasmalogen biosynthesis value (PBV) was calculated for each participant utilizing five ratios of four key glycerophospholipids. A linear regression model of global cognition was constructed with PBV, adjusted for sex, age, education, total cholesterol, and body mass index. In participants with 3T MRI brain imaging, the association between PBV and white matter hyperintensities (WMH) was explored.

**RESULTS:** Of the 298 participants, the mean age was 74.6 years, mean education was 15.6 years, and 84% were women. The median PBV was 0.42 (interquartile range: 0.22 to 1.14). A unit higher in PBV was suggestively associated with a 0.17 β-unit higher cognitive z-score (SE =0.09, p = 0.06). In 254 participants with MRI data, an increase in log_10_ SD of PBV suggested the less white matter hyperintensities (estimate = –0.20, SE = 0.12, p = 0.08).

**DISCUSSION:** In older African Americans, higher PBV was associated with higher level of global cognition, and potentially lower levels of brain white matter hyperintensities. Larger studies are needed in additional cohorts to determine if PBV is associated with annual rate of change in cognitive function.

**KEY POINTS:** *What was known before this study?:* Recent studies, in primarily older, white participants have found increased plasmalogen levels to be associated with better cognition. Given the prevalence of lipid related chronic conditions and disproportionate burden of cognitive impairment in African Americans, we explored whether plasmalogen ratios may be associated with brain function and structure in this population.

*What did we learn in this analysis?:* Utilizing data from 298 participants recruited from the Minority Aging Research Study (MARS) and Rush Clinical Core, we tested whether higher blood plasmalogen levels were associated with (1) lowered cognitive scores, and (2) increased white matter hyperintensities (WMH) on brain imaging. We found that higher PBV values showed a trend towards better global cognition. We found that PBV was associated with WMH.

*What do we still have to learn?:* Future studies may need to investigate these novel biomarkers and their neuroprotective capabilities in persons with cognitive impairment or in later stages of cognitive decline in addition to their longitudinal fluctuations to better gauge their role in dementia and AD.

## INTRODUCTION

As of 2024, the CDC reported that 13.9% of the United States’ population over age 55 experienced cognitive impairment, with researchers estimating a lifetime risk of dementia being 42% (Doctrow, 2025). Recent studies suggest that African Americans have twice the likelihood of developing dementia when compared to other racial and ethnic groups, having more risk factors, greater cognitive deficits, and increased neuropsychiatric symptoms (Lennon et al, 2021; Cielsa et al, 2024). Several explanations have been proposed, including higher presence of cardiovascular risk factors and diseases, differences in years and quality of education, and unequal access to healthcare. However, none of these listed factors can fully explain this disparity, suggesting additional underlying novel factors. Lipid metabolism is a potentially modifiable biomarker that has been examined in predominantly White populations but has received little attention in Black populations. Given the prevalence of chronic conditions within this demographic that involve lipid metabolism (e.g. cardiovascular disease, diabetes), understanding how lipid profiles are associated with cognition may lead to a deeper understanding of brain health, and ultimately brain health disparities.

Lipids are diverse molecules that have many different functions in the body including homeostasis, neurotransmission, and transporting materials within the cellular environment. The brain is a lipid-rich environment with dry brain weight comprising 50% of cerebral lipids and roughly 20% phospholipid mass is associated with a particular class of glycerophospholipids called ether phospholipids (Braverman and Moser, 2012; Kao et al, 2015; Farooqui et al, 2000; Dorninger et al, 2015). Lipid health is critical to the structural and physiological functions of the brain (Honsho et al, 2015). Recent studies have delved into the role of lipid homeostasis in AD pathology. The most abundant subclass of ether phospholipids are plasmalogens, whose biosynthesis is initiated by peroxisomes and completed in the endoplasmic reticulum. Plasmalogens are a subclass of lipids found in cell membranes and enriched in polyunsaturated fatty acids (PUFAs). They play an essential role in cellular membrane composition and dynamics. For instance, in the brain, plasmalogens are more concentrated within lipid rafts, which are microdomains in the cell membrane, and support signal transduction between cells. Plasmalogens also play an important role in intracellular cholesterol transport and esterification (Dorninger et al, 2015; Honsho et al, 2015). Over the past two decades, an increasing number of studies have examined plasmalogen levels as a target for healthy aging. Older people living with Alzheimer’s Disease have significantly lower levels of plasmalogens as compared to their age-matched controls (Goodenowe and Senanayake, 2019). However, most of the research has been done primarily within white racial/ethnic cohorts with varying levels of cognitive function from normal to within the dementia range. Little is known about whether the association exists within other racial/ethnic cohorts.

**Figure 1.**
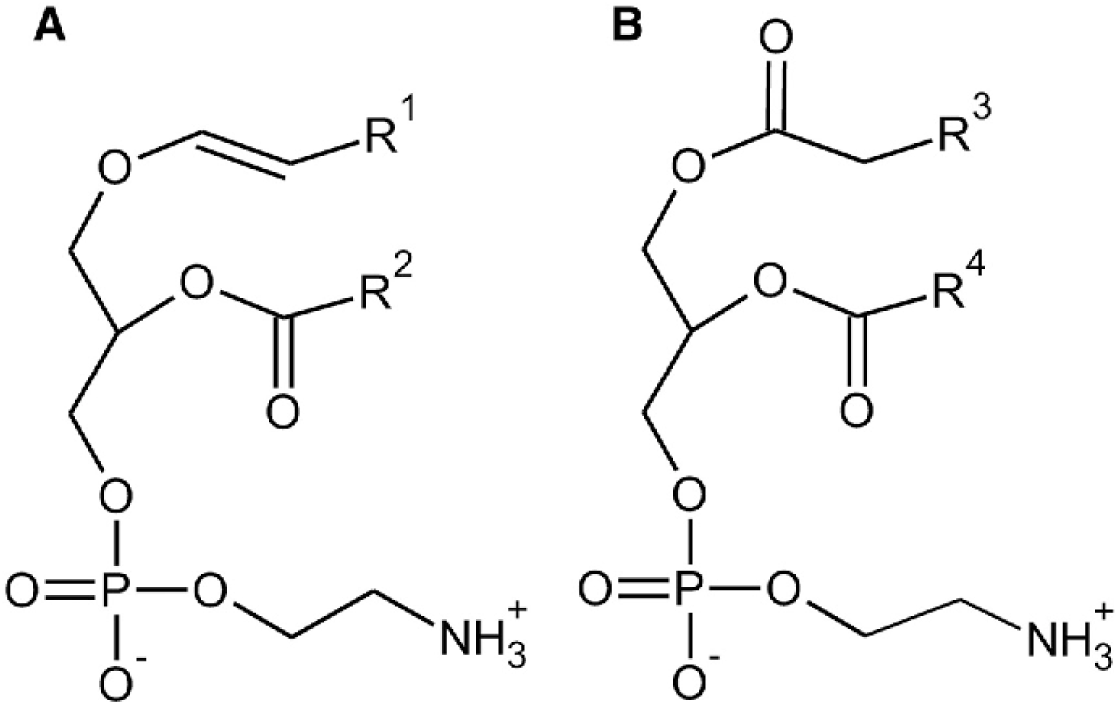
Plasmalogen Structure. (A) shows the chemical structure of plasmalogen. It is characterized by a vinyl ether (alkenyl ether) linkage at the sn-1 position of the glycerol backbone. (B) shows the structure of phosphatidylethanolamine, in which fatty acyl chains are ester-linked to the glycerol backbone; the sn-1 position contains an ester carbonyl with a double-bonded oxygen (C=O), rather than an ether linkage.

The overall goals of this study were to (1) measure a panel of plasmalogens and related phospholipids in stored blood of participants from two cohort studies of aging and to (2) test whether plasmalogens and related phospholipids are associated with key markers of brain function and structure. The underlying hypotheses were that higher plasmalogen levels were related to (1) a higher level of cognitive function and (2) lowered volume of white matter hyperintensities (WMH) within community dwelling, older African Americans.

## METHODS

### Cohort Participant Source

Participants were enrolled in the Minority Aging Research Study (MARS) or the Rush Clinical Core of the Rush Alzheimer’s Disease Center. For each study, interested participants provided a written informed consent which had been approved by an Institutional Review Board at Rush University Medical Center. MARS started in 2004 and consists of more than 850 community-dwelling African Americans; the Rush Clinical Core started in 2008, consisting of more than 300 community-dwelling African Americans. Participants from both cohorts were recruited from churches, senior buildings, and organizations that cater to older African Americans. Participation eligibility in either study required an individual to self-identify as an African American/Black, be 65 years of age or older, and have self-reported as having no known dementia diagnosis at the time of enrollment. Dementia is evaluated during yearly visits based on the criteria of the Joint Working Group of The National Institute of Neurological and Communicative Disorders and Stroke and the Alzheimer’s Disease and Related Disorders Association (Arvanitakis et al, 2015). Eligibility criteria for the current analysis included: 1) self-identification as African American, 2) a non-missing score for cognitive function, 3) presence of a serum sample concurrent to cognitive function testing, and 4) no dementia cognitive impairment at time of visit.

### Clinical, Blood Sample, and Brain Imaging Procedures

#### Clinical

Participants from both cohorts agreed to annual clinical evaluations. Data collection took place in the participants’ home, consisting of a review of their medical history, a comprehensive neuropsychological test battery to assess cognitive function, and an assessment of a wide range of environmental, sociocultural, behavioral, and biological risk factors. During their yearly assessment, participants had a blood draw and were invited to have a brain scan every other year if they were interested and eligible.

#### Brain Imaging

Brain MRI was conducted on a 3 Tesla General Electric MRI scanner to obtain high-resolution T 1-weighted anatomical data and T 2-weighted fluid-attenuated inversion recovery (FLAIR) data which are automatically segmented to generate total WMH volume data using a deep learning approach, as recently described elsewhere (Arvanitakis et al, 2015). Imaging corrected for offset due to scanner variability by adding a scanner variable in the model of the statistical analysis.

#### Blood samples

Consent was obtained as part of MARS and the Rush Clinical Core to collect and store blood samples for future analysis. Serum samples from participants were diluted and directly analyzed using ThermoFisher Scientific LTQ Orbitrap mass spectrometer in positive and negative electrospray ionization modes. A common pooled reference sample was used as quality control as described before (Goodenowe et al, 2007). Phospholipid peaks were identified based on accurate mass as previously described (Goodenowe et al, 2022). The characteristic fatty acids of lipidomic species of interest were measured using the negative electrospray ionization mass spectrometry.

### Outcome Variables

Cognitive data and MRI scans available proximate to collection of the serum sample for lipidomic measurement were utilized to measure cognitive function and WMH as the two outcome variables of interest.

#### Cognitive Function

Each study included a uniform structured clinical evaluation consisting of a battery of 19 tests to document level of cognition. The results of these tests were then used to create a composite measure of global cognition based on the average z-score using the means and standard deviations from the baseline visit (Bennett et al, 2012; Hagve and Christopherson, 1986). The results of these tests were also stratified to summarize cognition into five domains: episodic memory (Logical Memory Ia, Logical Memory IIa, Immediate story recall, Delayed story recall, Word List Memory, Word List Recall, Word List Recognition), semantic memory (Boston Naming Test, Category Fluency, Wide Range Achievement Test), working memory (Digit Span Forward, Digit Span Backward, Digit Ordering), perceptual speed (Symbol Digit Modalities Test, Number Comparison, Stroop word reading, Stroop color naming), and visuospatial ability (Judgment of Line Orientation and Standard Progressive Matrices). Only data from the assessment proximate to the serum sample collection were used for the current study.

#### Brain Structure

The total volume of brain tissue affected by WMH is measured for each participant and then is divided by the corresponding intracranial volume. Logarithmically transformed total WMH volumes were used for analysis.

### Blood Plasmalogen Variables

Phosphatidyl ethanolamine 16:0/22:6 (PE226) and three individual ethanolamine plasmalogens [16:0/22:6 (PL226), 16:0/22:4 (PL224), and 18:0/20:5(PL205)], each consisting of a total of 38 carbons in the combination of their sn-1 and sn-2 positions, were identified based upon their sn-2 fatty acid side chains. Each phospholipid level was divided by its sex-specific mean and then log_10_ transformed. Five ratios were then calculated with the 4 transformed phospholipid levels: PL205/PE226, PL226/PE226, PL205/PL224, PL226/PL224, and PL205/PL226. A composite plasmalogen biosynthesis value (PBV) was created for each participant by averaging these five ratios.

### Other Covariates

Other covariates utilized within this analysis were sex, age at time of visit, and education, which were identified by participants through self-report questions at the time of entry to each study. Total cholesterol was measured in blood and body mass index (BMI) was calculated by dividing measured weight (kg) by self-reported height (m^2^).

### Statistical Analyses

At the analytic baseline, women and men participants were compared on demographic and lipidomic characteristics using Welch’s two-sample t-tests. Primary analyses used linear regression models to evaluate the association between PBV and global cognitive function, adjusting for sex, age, education, total cholesterol, and BMI. To further examine domain-specific relationships, each of the five lipid ratios composing the sex-adjusted PBV was tested as a predictor in separate linear regression models for global cognition and for the five cognitive domains: episodic memory, visuospatial ability, perceptual speed, semantic memory, and working memory. PBV and lipid ratios were modeled on their original scales; for interpretability, we additionally report effects per 1 SD increase by multiplying the estimated coefficient by the sample SD of the predictor. Secondary analysis was conducted in a subset of participants with 3T MRI brain imaging. Similarly adjusted linear models were used to evaluate the relationship between PBV or lipid ratios and log_10_-transformed WMH volume. All WMH models additionally adjusted for scanner. To evaluate whether PBV and WMH were independently associated with global cognition, models were fit to PBV alone, WMH alone, and both PBV and WMH jointly. To explore effect modification, an additional PBV×WMH interaction term was tested. All models were stratified by sex to explore potential differences in associations, and regression coefficients (β) were estimated to aid interpretability and cross-comparison. Given the exploratory nature of this pilot analysis, associations with p < 0.05 were considered statistically significant, while associations with 0.05 ≤ p < 0.10 were considered suggestive. No corrections were made for multiple comparisons. All analyses were conducted in R version 4.4.1 (2024-06-14 ucrt).

## RESULTS

### Participant Characteristics

1,386 MARS and Rush Clinical Core participants were screened for having a concurrent serum sample for lipid analysis along with cognitive function testing and no cognitive impairment and 298 individuals meeting this criterion were randomly selected. Of these 298 participants, the mean age was 74.6 years, mean education was 15.6 years, and 84% were women. The median baseline PBV serum ratio was found to be 0.42 (interquartile range: 0.22 to 1.14). Analysis of these key phospholipids and their ratios were further broken down to identify possible changes across sex-specific means (see table 1). The median z-score of global cognitive function was 0.36 for women (SD = 0.45) and 0.26 for men (SD = 1.60).

**Table 1.** Baseline analytical demographic and phospholipid values of women and men African Americans in cross sectional analysis. Abbreviations: PBV = Plasmalogen Biosynthesis Value; BMI = Body Mass Index; WMH = White Matter Hyperintensity.

| Demographics |  |  |  |  |  |
| --- | --- | --- | --- | --- | --- |
|  | Women (n =250) |  | Men (n=48) |  |  |
| Variable | Mean | SD | Mean | SD | p- value |
| Age (years) | 74.58 | 6.09 | 74.45 | 5.74 | 0.89 |
| Total Cholesterol (mg/dL) | 189.16 | 0.22 | 169.91 | 1.26 | 0.01 |
| BMI (w/h<sup>2</sup>) | 29.21 | 0.26 | 28.65 | 0.16 | 0.32 |
| Education (years) | 15.58 | 3.28 | 15.42 | 3.08 | 0.74 |
| Global Cognition | 0.36 | 0.45 | 0.26 | 1.60 | 0.16 |
| WMH volume | -0.35 | 0.49 | -0.45 | 0.53 | 0.26 |
| Serum Ethanolamine Glycerophospholipids |  |  |  |  |  |
| Variable | Mean | SD | Mean | SD | p- value |
| PE226 | 8.31 | 0.49 | 8.04 | 0.53 | 0.00 |
| PL224 | 9.34 | 0.28 | 9.29 | 0.30 | 0.23 |
| PL205 | 9.22 | 0.28 | 9.13 | 0.32 | 0.07 |
| PL226 | 8.65 | 0.42 | 8.51 | 0.43 | 0.04 |
| Plasmalogen Ratios |  |  |  |  |  |
| Variable | Mean | SD | Mean | SD | p- value |
| PBV | -0.38 | 0.26 | -0.16 | 0.32 | 0.00 |
| PL226/PE226 | 4.46 | 6.64 | 7.35 | 9.77 | 0.05 |
| PL205/PE226 | 16.11 | 19.22 | 27.95 | 35.39 | 0.03 |
| PL205/PL226 | 0.32 | 0.23 | 0.30 | 0.20 | 0.43 |
| PL226/PL224 | 0.29 | 0.33 | 0.24 | 0.24 | 0.26 |
| PL205/PL224 | 0.84 | 0.84 | 0.74 | 0.30 | 0.16 |

### Association of Plasmalogen and Global Cognitive Function

In the full analytic cohort of 298 participants, a unit higher in PBV was suggestively associated with a 0.17 β-unit higher cognitive z-score (SE =0.09, p = 0.06) after adjustment for demographic and clinical covariates. As PBV was significantly different for women as compared to men, adjusted models stratified by sex were created. This unit higher suggestive association with PBV was also found in women (estimate = 0.17, SE = 0.10, p = 0.096), though no association was found for men even with the parameter estimate being the same (estimate = 0.17, SE = 0.19, p = 0.38).

Linear regression models were conducted for the overall cohort to examine the plasmalogen ratios with global cognitive score and the 5 cognitive domains for the overall cohort. An association was suggested between a SD higher in the PL226/PL224 and global cognition (estimate =0.16, SE =0.07, p=0.03) as well as for PL205/PL226 ratio and global cognition (estimate = 0.20, SE = 0.11, p=0.06) (see Supplemental Figure 1). These analyses were then conducted separately for women and men to determine if there were within sex differences (see Figure 1). For women, a higher global cognitive score was associated with a SD increase in the PL226/PL224 ratio (estimate = 0.16, SE = 0.08, p = 0.04) and with a SD increase in the PL205/PL226 ratio (estimate = 0.23, SE = 0.12, p = 0.05). The greatest impact of these two ratios was for working memory (see Figure 1). No significant associations were found for each of the 5 plasmalogen ratios that constitute PBV and global cognition in men (see Figure 1).

**Figure 1.**
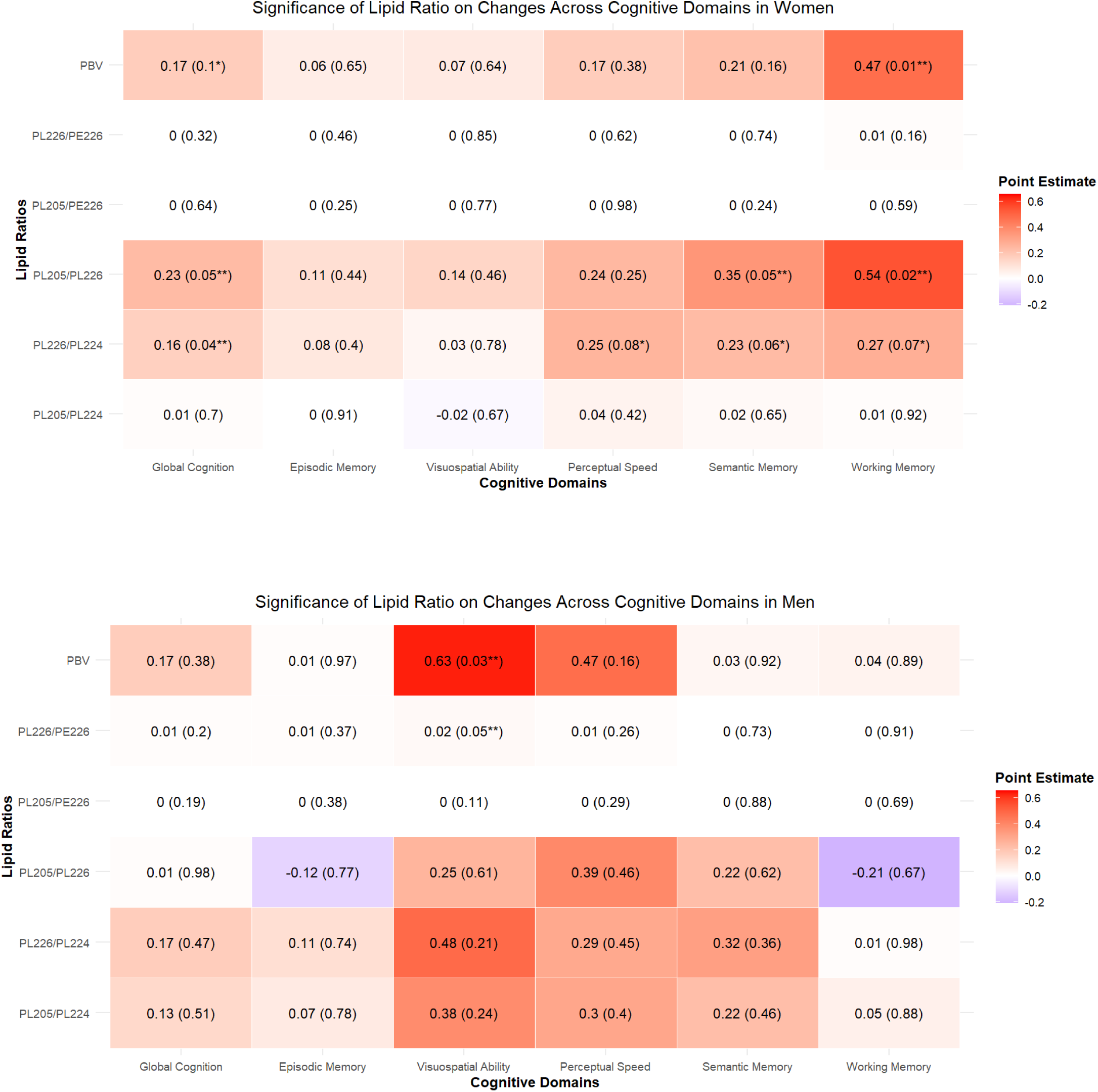
Heat map of point estimates of linear regression models for all participants with cognitive testing available, separated by sex. Labeled as estimate(p-value) with p-values labeled p≤0.05 “**”, and <0.10 “*” for clarity. Abbreviations: PBV = Plasmalogen Biosynthesis Value.

### Association of Plasmalogen and White Matter Hyperintensities on Brain Imaging

A subset of 254 participants of the full analytic cohort with available 3T MRI data within a two-year window surrounding the corresponding blood draw date were included for secondary MRI analysis. Of this subset, the median z-score for WMH volume was –0.35 (SD = 0.49) for women and –0.45 (SD = 0.53) for men. The analysis of PBV and WMH volume in the subset of participants with MRI suggests a potential physiological mechanism (estimate = –0.20, SE = 0.12, p = 0.08) that is not seen in sex-specific models (see table 2). In addition, we found little evidence that the association between PBV and cognition differed by WMH burden based on the PBV×WMH interaction term (see Models 3 and 4 in supplemental table 1).

**Table 2.** Linear Regression Model of Normalized WMH Volume and Lipidomic Ratios. Abbreviations: PBV = Plasmalogen Biosynthesis Value.

| Variable | Total MRI Subset n=254<br>Estimate (SE, p-value) | Women n=217<br>Estimate (SE, p-value) | Men n=37<br>Estimate (SE, p-value) |
| --- | --- | --- | --- |
| PBV | -0.20 (0.12, 0.08) | -0.16 (0.13, 0.21) | -0.40 (0.26, 0.13) |
| PL205/PE226 | 0.00 (0.00, 0.22) | 0.00 (0.00, 0.52) | 0.00 (0.00, 0.51) |
| PL226/PE226 | -0.01 (0.00, 0.10) | -0.01 (0.00, 0.25) | -0.01 (0.01, 0.40) |
| PL205/PL224 | -0.01 (0.04, 0.71) | -0.01 (0.04, 0.72) | -0.14 (0.27, 0.59) |
| PL226/PL224 | -0.14 (0.09, 0.11) | -0.15 (0.09, 0.12) | -0.14 (0.30, 0.64) |
| PL205/PL226 | -0.17 (0.12, 0.20) | -0.14 (0.14, 0.31) | -0.41 (0.40, 0.32) |

## DISCUSSION

Cross-sectional analysis of data from 298 older, community-dwelling African Americans found a positive association between higher PBV, a composite measure of important plasmalogen ratios, and better brain function as measured by global cognitive function level. When PBV value was investigated with relative Z score of WMH volume for a subset of 254 participants, the parameter estimate showed a higher PBV was associated with lower WMH volume.

Previous studies using cross-sectional lipidomic data in primarily white populations of varying cognitive impairment from normal cognition to dementia have found that higher serum plasmalogen levels are correlated with a higher global cognitive function level. In work done with measures of PBV in the Rush Memory and Aging Project (MAP) with biomarker and cognitive data on 1,205 participants, PBV was associated with better function across all cognitive domains with its strongest effects on episodic memory. The parameter estimate for the association in the MAP participants was almost double what was seen in this study (Goodenowe and Senanayake, 2019). This difference may be due to the large variability of cognitive impairment that was included within that study. In this analysis, only participants with no clinical impairment at the time of blood draw were included. Additionally, both the MAP and MARS studies showed significant differences between male and female participants, with this study finding a greater effect of PBV on working memory followed by perceptual speed and semantic memory in females. In a 2023 ADNI study on WMH progression on cognition, sex differences were found in both risk factors and location of WMH burden, with females having increased progression in total, frontal, and deep white matter and males having increased progression in occipital white matter (Morrison and Collins, 2023), corresponding to the cognitive results listed in this study. A similar lipidomic study was done in the ADNI study that pointed to some molecules being associated with imaging changes (Nho et al, 2021; Kling et al, 2020), but a direct comparison to the lipidomic ratios used for PBV could not be determined. Unlike prior studies, we evaluated whether WMH burden accounted for the PBV–cognition association. When PBV and WMH were included in the same model, the PBV estimate was not attenuated in the overall cohort or within each sex.

In this study, five composite ratios were combined to create the composite PBV and may give some mechanistic insights. Two of the ratios, PL226/PE226 and PL205/PE226, assess the relative plasmalogen ethanolamine and phosphatidyl ethanolamine pool size. By reflecting the balance between ether-linked and ester-linked ethanolamine species, these two ratios serve as a marker of plasmalogen homeostasis relative to total phosphatidyl ethanolamine (Olazarán et al, 2015; Abdullah et al, 2017). The remaining three ratios (PL226/PL224, PL205/PL224, and PL205/PL226) reflect differences in plasmalogen acyl-chain composition and are influenced by multiple lipid-metabolic pathways, including peroxisomal metabolism. As PL205/PL226 and PL226/PL224 are associated with global cognition within women but not within men, further studies with larger sample sizes are needed to elucidate the underlying biological mechanisms and potential for within sex differences. Finding PBV does not significantly impact the influence of WMH changes on global cognition suggests a process for the mechanism of plasmalogens that is not vascular mediated in nature.

A strength of this analysis is the well-characterized cohort of older, community-dwelling African Americans that underwent similar study procedures, blood collection and processing, and imaging studies as the prior work in the Rush Memory and Aging Project. In addition, assays for the lipidomic markers were done by the same laboratory leadership group. Finally, we were able to obtain consistent brain structure data through an MRI protocol and quantitative deep learning model to generate the percentage of WMH.

Limitations of the study included cross-sectional design which does not enable examination of features associated with progression. Second, samples for this analysis were taken specifically from individuals diagnosed as having no cognitive impairment while previous studies have taken cross-sectional data from individuals within a wide range of cognitive functions, including mild cognitive impairment and dementia. There is the possibility that the biochemical changes previously identified may be indicative of later AD progression and decreased cognition.

In conclusion, we found that higher levels of PBV, a composite plasmalogen ratio, was associated with higher global cognitive function; however, the interaction of PBV and WMH changes is not suggestive of a solely vascular mechanism for plasmalogen influence on cognition. As this analysis was conducted in well-characterized, older community-dwelling, African American women and men, our work advances the understanding of lipidomics in relation to cognition and aging. However, future studies may need to investigate these novel biomarkers and their neuroprotective capabilities in persons with cognitive impairment or in later stages of cognitive decline in addition to their longitudinal fluctuations to better gauge their role in dementia and AD.

## DATA AVAILABILITY STATEMENT

The raw data supporting the conclusions of this article will be made available by the authors. Requests for data can be submitted at https://www.radc.rush.edu/

## ETHICS STATEMENT

This study was completed in accordance with the Helsinki Declaration and was reviewed and approved by a Rush University Medical Center Institutional Review Board.

## AUTHOR CONTRIBUTIONS

AAW was involved in the design, data collection, analysis of data and drafting with critical review of the manuscript. RCS was involved in the design, data collection, analysis of data, and drafting with critical review of the manuscript. DBG was involved in the design, analysis of data and specimens, and drafting with critical review of the manuscript. LLB was involved in design, data and specimen collection, analysis of data, and drafting with critical review of the manuscript. LD and VS were involved in critical review of the manuscript.

---

**Supplementary Figure 1.**
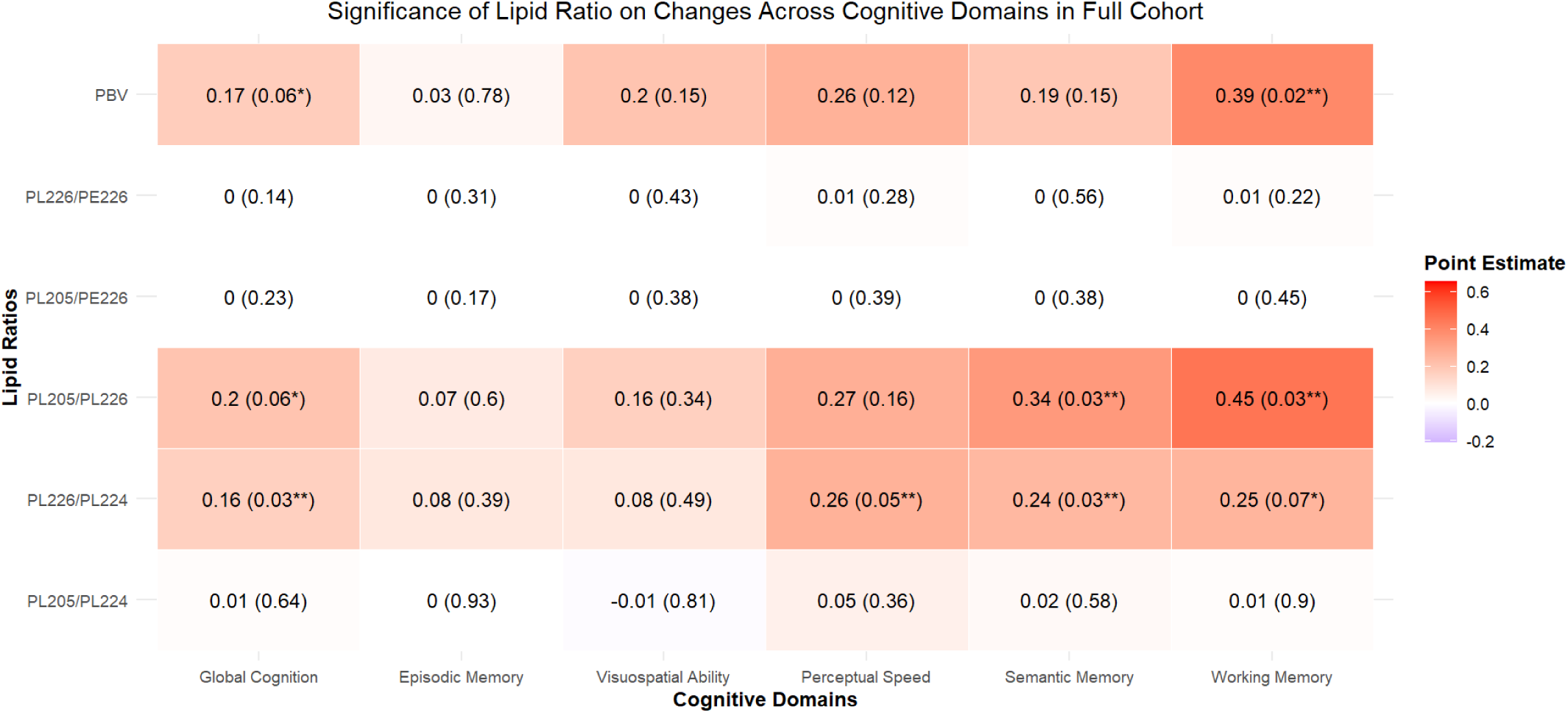
Heat map of point estimates of linear regression models for all participants with cognitive testing available. Labeled as estimate(p-value) with p-values labeled p≤0.05 “**”, and <0.10 “*” for clarity. Abbreviations: PBV = Plasmalogen Biosynthesis Value.

**Supplementary Table 1.**
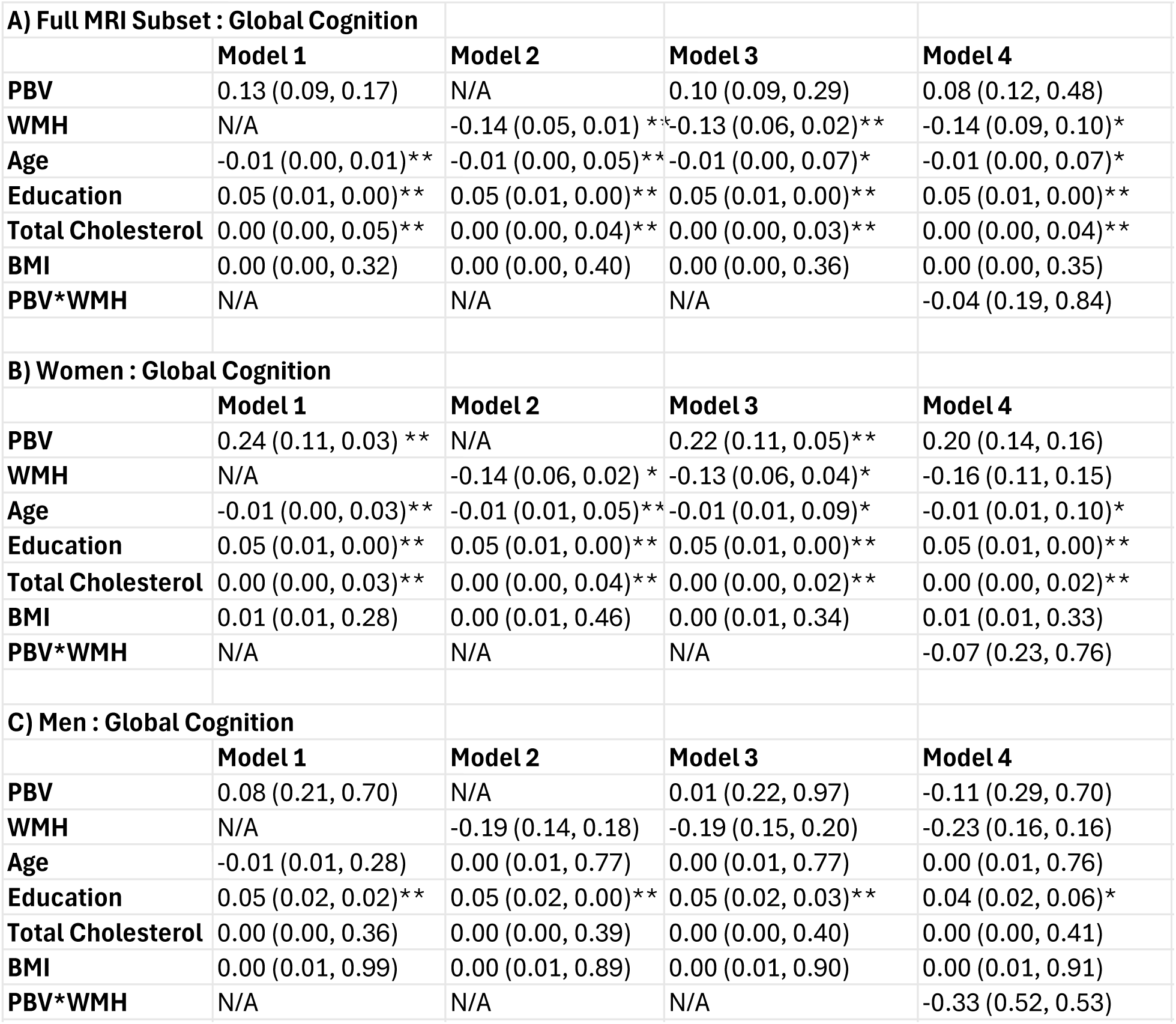
Linear Regression Models of Lipidomic Ratios on Cognitive Scoring in Women and Men with MRI available (n=254). Model 1 includes PBV with other covariates; Model 2 includes WMH with covariates; Model 3 includes PBV and WMH along with covariates; Model 4 includes PBV, WMH, and the PBV*WMH interaction along with same covariates. P values are labeled p≤0.05 “**”, and ≤0.10 “*” for clarity. Abbreviations: PBV = Plasmalogen Biosynthesis Value; BMI = Body Mass Index; WMH = White Matter Hyperintensity.

